# Altered EEG markers of synaptic plasticity in a human model of NMDA receptor deficiency: anti-NMDA receptor encephalitis

**DOI:** 10.1101/2020.09.28.20203265

**Authors:** Silvano R. Gefferie, Angelina Maric, Hanne Critelli, Sophie Gueden, Gerhard Kurlemann, Salome Kurth, Margherita Nosadini, Barbara Plecko, Maya Ringli, Kevin Rostásy, Stefano Sartori, Bernhard Schmitt, Agnese Suppiej, Patrick Van Bogaert, Flavia M. Wehrle, Reto Huber, Bigna K. Bölsterli

**Affiliations:** Department of Neuropediatrics and Children’s Research Center, University Children’s Hospital Zurich, 8032, Zurich, Switzerland; Stichting Epilepsie Instellingen Nederland (SEIN), 2103 SW, Heemstede, The Netherlands; Department of Neurology, University Hospital Zurich, University of Zurich, 8091, Zurich, Switzerland; Service de Pédiatrie, CHU d’Angers, 49933, Angers, France; Children’s Hospital Bonifatius Klinik, 49808, Lingen, Germany; Pulmonary Clinic, University Hospital Zurich, 8091, Zurich, Switzerland; Department of Psychology, University of Fribourg, 1700, Fribourg, Switzerland; Pediatric Neurology and Neurophysiology Unit, Department of Women’s and Children’s Health, University of Padua, 35122, Padua, Italy; Department of Pediatrics and Adolescent Medicine, Division of General Pediatrics, Medical University of Graz, 8036, Graz, Austria; Department of Neurology, Inselspital, University Hospital Bern, 3010, Bern, Switzerland; Department of Pediatric Neurology, Children’s Hospital Datteln, Witten/Herdecke University, 58448, Datteln/Witten, Germany; Department of Medical Sciences, Pediatric Section, University of Ferrara, 44121, Ferrara, Italy; Child Development Center, University Children’s Hospital Zurich, 8032, Zurich, Switzerland; Department of Neonatology and Pediatric Intensive Care, University Children’s Hospital Zurich, 8032, Zurich, Switzerland; Department of Child and Adolescent Psychiatry and Psychotherapy, Psychiatric Hospital, University of Zurich, 8091, Zurich, Switzerland

**Keywords:** *NMDA receptor*, *encephalitis*, *synaptic plasticity*, *slow waves*, *sleep homeostasis*.

## Abstract

Plasticity of synaptic strength and density is a vital mechanism enabling memory consolidation, learning, and neurodevelopment. It is strongly dependent on the intact function of N-methyl-D-aspartate receptors (NMDAR). The importance of NMDAR is further evident as their dysfunction is involved in many diseases such as schizophrenia, Alzheimer’s disease, neurodevelopmental disorders, and epilepsies. Synaptic plasticity is thought to be reflected by changes of sleep slow wave slopes across the night, namely higher slopes after wakefulness at the beginning of sleep than after a night of sleep. Hence, a functional NMDAR deficiency should theoretically lead to altered overnight changes of slow wave slopes. Here we investigated whether pediatric patients with anti-NMDAR encephalitis, being a very rare but unique human model of NMDAR deficiency due to autoantibodies against receptor subunits, indeed show alterations in this sleep EEG marker for synaptic plasticity.

We retrospectively analyzed 12 whole-night EEGs of 9 patients (age 4.3-20.8 years, 7 females) and compared them to a control group of 45 healthy individuals with the same age distribution. Slow wave slopes were calculated for the first and last hour of non-rapid eye movement (NREM) sleep (factor ‘hour’) for patients and controls (factor ‘group’). There was a significant interaction between ‘hour’ and ‘group’ (*p* = 0.013), with patients showing a smaller overnight decrease of slow wave slopes than controls. Moreover, we found smaller slopes during the first hour in patients (*p* = 0.022), whereas there was no group difference during the last hour of NREM sleep (*p* = 0.980). Importantly, the distribution of sleep stages was not different between the groups, and in our main analyses of patients without severe disturbance of sleep architecture, neither was the incidence of slow waves. These possible confounders could therefore not account for the differences in the slow wave slope values, which we also saw in the analysis of the whole sample of EEGs.These results suggest that quantitative EEG analysis of slow wave characteristics may reveal impaired synaptic plasticity in patients with anti-NMDAR encephalitis, a human model of functional NMDAR deficiency. Thus, in the future, the changes of sleep slow wave slopes may contribute to the development of electrophysiological biomarkers of functional NMDAR deficiency and synaptic plasticity in general.

**Highlights:**

- Changes of slow waves in overnight EEGs are thought to reflect synaptic plasticity.
- Synaptic plasticity is strongly dependent on intact NMDAR function.
- Antibody-mediated NMDAR deficiency occurs in patients with anti-NMDAR encephalitis.
- In this human model of NMDAR deficiency, we found altered slow wave changes.
- Sleep EEG measures may mark NMDAR-related impairments of synaptic plasticity.

## 1. Introduction

N-methyl-D-aspartate receptors (NMDAR) are a class of ionotropic glutamate receptors ubiquitously distributed throughout the central nervous system. Due to their importance, it is not surprising that a dysfunction of NMDAR is involved in many diseases such as schizophrenia (Tsai and Coyle, 2002), Alzheimer’s disease (Zhang et al., 2016), and epilepsies and neurodevelopmental disorders (Burnashev and Szepetowski, 2015). NMDAR are primarily known for their pivotal role in long-term synaptic plasticity (Dupuis et al., 2014; Escobar et al., 1998). Synaptic plasticity involves behavior-dependent synaptic potentiation during wakefulness, being the basic mechanism of learning, and behavior-independent renormalization of synaptic strength and density during sleep, which has been hypothesized to ensure synaptic homeostasis (Tononi and Cirelli, 2014). Various studies suggested that the processes that underlie synaptic plasticity and homeostasis, i.e. changes in net synaptic strength and synaptic density, are reflected by changes of sleep slow wave (0.5 – 4Hz) characteristics measured in the surface EEG (Esser et al., 2007; Riedner et al., 2007; Vyazovskiy et al., 2009).

Slow waves stem from (near) synchronous activity of large populations of cortical neurons (Esser et al., 2007; Vyazovskiy et al., 2009), which has been related to the strength of synaptic connections (Esser et al., 2007). They are most prominent in non-rapid eye movement (NREM) sleep and can be characterized by various parameters such as incidence, power, amplitude, and slope (Esser et al., 2007). Of these, the slope has been suggested to be the preferred EEG marker for cortical synchrony, being the closest reflection of synaptic strength (Riedner et al., 2007; Vyazovskiy et al., 2009). Accordingly, changes of slow wave slopes from the first hour (FH) to the last hour (LH) of NREM sleep have been used to indirectly measure synaptic plasticity (Bölsterli et al., 2011; Fattinger et al., 2015). This typically results in steeper slopes of slow waves at the beginning of sleep because of synaptic potentiation during previous wakefulness and a homeostatic decrease of slow wave slopes across the night (Riedner et al., 2007).

The analysis of sleep slow waves has indicated that synaptic plasticity may be challenged in various conditions, including major depressive disorder (Armitage et al., 2000), encephalopathy with status epilepticus during sleep (ESES) (Bölsterli et al., 2011) and infantile spasms (Fattinger et al., 2015). Since synaptic plasticity requires properly functioning NMDAR, a dysfunction would expectably result in altered changes of slow wave characteristics during a night’s sleep. Such a functional NMDAR deficiency may emerge in different ways, for example by interventions such as gene editing and receptor antagonism (Campbell and Feinberg, 1996; Miyamoto et al., 2003), or by antibodies against the NMDAR as seen in anti-NMDAR encephalitis.

Anti-NMDAR encephalitis is a well-recognized but very rare disease due to autoantibodies against the NR1-subunit of NMDAR (Dalmau et al., 2008, 2007). Anti-NMDAR antibodies disturb receptor functionality by cross-linking them, which leads to their internalization into neurons, causing a functional NMDAR deficiency (Hughes et al., 2010). Accordingly, such patients constitute a human model of NMDAR deficiency, which suggests that the receptors’ role in synaptic plasticity is impaired.

In this retrospective, multicenter, case-control study, we hypothesized that the changes of slow wave slopes across the night in pediatric patients with anti-NMDAR encephalitis, representing a human model of NMDAR deficiency, are altered, i.e. show diminished patterns of change. To this end, we compared changes of slow wave slopes from the FH to the LH of NREM sleep between patients and healthy controls of corresponding age. To rule out that differences in other sleep variables such as sleep stage distributions or the mere counts of slow waves would drive group differences in slow wave slopes, we additionally compared these variables between the groups.

## 2. Materials and Methods

### 2.1. Participants

Patients diagnosed with anti-NMDAR encephalitis were provided by the Department of Neuropediatrics, University Children’s Hospital Zurich (Switzerland); the Department for Women’s and Children’s Health, Pediatric Neurology and Neurophysiology Unit, University of Padua (Italy); the Department of Pediatric Neurology, Children’s Hospital Datteln, University Witten/Herdecke (Germany); the University Hospital Muenster, Westfalian Wilhelms University (Germany); and the Service de Pédiatrie, CHU d’Angers (France). Twenty-one EEGs from thirteen patients (ten females) recorded between 2008 and 2019 were available. We included patients that met all the following inclusion criteria:

- Detection of anti-NMDAR antibodies in serum and/or cerebrospinal fluid
- At least one whole-night sleep-EEG recording of sufficing data quality and without highly pathological changes that allowed scoring of sleep stages
- Under the age of 21 years at the time of EEG recording

Nine EEGs from nine patients had to be excluded because of the following reasons: inability to score sleep stages, for example due to excessive artifacts (six EEGs); the EEG not being a whole-night registration (one EEG); incompleteness of the EEG (one EEG); and a lack of sleep during the recording (one EEG). Consequently, we included 12 whole-night EEGs from 9 patients (7 females, age 4.3 – 20.8 years, mean 12.3 years) at different stages of their disease. Clinical and electrographic characteristics of included patients are provided in Table 1 and Supplementary Table 1.

**Table 1.** Clinical and electrographic characteristics of anti-NMDAR encephalitis patients at the time of EEG registration.

| Patient<br>(#) | Sex | EEG | Age<br>(years) | Time since diagnosis | Clinical presentation | Sleep EEG<br>structure<br>anomalies |
| --- | --- | --- | --- | --- | --- | --- |
| 1 | f | A^ | 14.7 | ≅3mt | Started to speak 2d before EEG, speech problems (single words expressed, slurred pronunciation).<br>Child-like behaviour. Fluctuating confusion, disorientation, agitation. Restlessness. Improving<br>flaccid paresis of the legs (critical illness myopathy, 75d in the intensive care unit) | Severe sleep<br>fragmentation |
|  |  | B | 14.9 | ≅5 ½mt | Clear clinical improvement compared to EEG A. Increased alertness, speaks more, still slightly<br>blurred and word-finding problems, better motor skills (sitting, walking some steps possible).<br>Behavioral inflexibility | None |
| 2 | m | A | 7.4 | <1mt | The morning before EEG, focal seizure with clonic jerking of the right-side of mouth. Mood<br>swings and slurred speech | None |
|  |  | B | 7.4 | <1mt | Clinical improvement compared to EEG A. Slight truncal ataxia (abnormal line gait with closed | Period (2.81hrs) |
|  |  |  |  |  | eyes). Emotional lability. Good visuoconstructive performance. Language impairment (pronunciation, word-finding, speech planning, vowel differentiation, and auditory memory; but normal comprehension), reduced memory capacity and concentration | of continuous wakefulness after the start of NREM sleep |
| 3 | f | A^ | 4.3 | $\cong 1 \frac{1}{2}$mt | Mutism. Interaction by eye contact preserved. Receptive language better preserved but limited to simple orders. Chorea, allowing walking with assistance and impairing fine motor skills. Neuropsychological testing impossible | Disrupted sleep architecture; frontal seizure patterns |
| 4 | f | A | 17.9 | $\cong 5$mt | No abnormalities | None |
| 5* | f | A | 20.8 | $\cong 63$mt after diagnosis,<br>48mt after relapse | No abnormalities | Period (3.24hrs) of continuous wakefulness after the start of NREM sleep |
| 6 | m | A | 13.0 | $\cong 1 \frac{1}{2}$mt | mRS 3. Can walk approximately ten steps without support. Starting oral feeding | None |
| | | B | 13.0 | $\cong 2$mt | mRS 3. Better sleep compared to EEG A. Echolalia. Hyperphagia. Intermittent verbal aggressiveness | None |
| 7 | f | A | 9.5 | ≅1mt | mRS 2. Speech difficulties (pronunciation and word finding). Fatigue | None |
| 8 | f | A^ | 9.7 | ≅1mt | Clinically, long sleep episodes. Eats and drinks in drowsiness, orofacial dyskinesia. Muscular hypotonia in extremities. Stable torso. Weak muscle tendon reflexes | Severe sleep fragmentation |
| 9 | f | A | 13.3 | Diagnosis made ≅3mt after EEG (the result of CSF specimen, taken at the time of EEG, received after 3mt) | Started to speak 2d before EEG, speech problems (single words expressed, slurred pronunciation). Child-like behaviour. Fluctuating confusion, disorientation, agitation. Restlessness. Improving flaccid paresis of the legs (critical illness myopathy, 75d in the intensive care unit) | None |

An EEG designated ‘A’ or ‘B’ indicates whether it concerns the first or, if applicable, the second available recording for the same patient, respectively.
^ EEGs not in the main statistical analysis (see 2. Materials and Methods for details).
\* Patient not included in the statistical analysis, but slow wave analysis results assessed individually (see 2. Materials and Methods for details).
d = day(s), hrs = hours, mRS = modified Rankin Scale, mt = month(s), NREM = non-rapid eye movement.

As representative references for each patient, we included five healthy controls with one whole-night EEG recording from studies conducted between 2008 and 2019 by the research group of Reto Huber from the University Children’s Hospital Zurich (Kurth et al., 2012; Wehrle et al., 2017; Volk et al., 2019; unpublished observations). Age-wise, these controls differed at most 1.1 years from the corresponding patient, resulting in an age distribution in controls (age 4.2 - 21.4 years, mean 12.4 years) that was comparable to that in patients. This procedure was carried out to avoid that age-related differences in sleep slow wave characteristics could eventually drive differences between patients and controls (Kurth et al., 2010b). Moreover, the sex of controls was matched for all but one patient. For this young male patient (7.4 years), we included three male and two female controls, as no more data of male controls in that age range were available. Importantly, sex influences on sleep parameters are thought to be minimal before puberty (Johnson et al., 2006; Knutson, 2005).

This study was performed at the University Children’s Hospital Zurich. All participants or their legal representatives gave written informed consent for the use of their non-identifiable, health-related data for research purposes – acknowledged by their local authorities. Ethical approval was provided by the ethical committee of the canton of Zurich (BASEC Nr. 2016-02154).

### 2.2. EEG recordings

The recordings from the patients in Zurich were performed using a Neurofile XP EEG System (Natus Europe GmbH, Munich, Germany), with 19 electrodes according to the international 10-20 system. Their EEG data were analogously band-pass filtered (0.016 – 300 Hz), saved at 256 Hz and then digitally band-pass filtered (0.53 – 70 Hz). At the Children’s Hospital Datteln, a Neurofax EEG-9210 System (Berger Medizintechnik GmbH, Gleisdorf, Österreich) was used. The signals were stored at 200 Hz and digitally band-pass filtered (0.53 – 70 Hz). At the Pediatric Clinical Hospital of Padua, EEG data were acquired with Galileo NT Software (EB Neuro S.p.A, Florence, Italy), analogously wide-band filtered (0 – 20 kHz) and consequently saved at 256 Hz. The EEGs from the University Hospital Angers were registered with a Deltamed Brainbox-1042 EEG amplifier (Natus Europe GmbH, Munich, Germany), stored at 256 Hz, and digitally high- and low-pass filtered at 0.17 Hz and 100 Hz, respectively. The EEGs from patients in Angers, Datteln and Padua had at least eight EEG electrodes placed on 10-20-positions. The six common electrodes among all EEGs were O1, O2, Fp1, Fp2 and temporal electrodes (either T3 and T4, or T5 and T6). The EEGs from Münster were excluded due to excessive artifacts.

To avoid inter-rater variability, all sleep stage scoring for patients was done by a single, trained EEG technician (H.C.) according to the standard criteria from the American Association of Sleep Medicine (AASM) (Iber et al., 2007), applied to 20-sec epochs.

EEGs for controls were recorded using a 128-channels high-density EEG system (Electrical Geodesics, Inc.; Sensor Net for long-term monitoring). The data were sampled at 500 Hz (analogously filtered between 0.01 and 200 Hz) and then digitally band-pass filtered (0.5 – 50 Hz). Scoring was done by investigators of the original studies (Kurth et al., 2012; Wehrle et al., 2017; Volk et al., 2019; unpublished observations) according to the guidelines from the AASM (Iber et al., 2007), applied to 20-sec epochs.

### 2.3. EEG analyzes

After acquisition and scoring of all patient and control EEG recordings, data were further analyzed in MATLAB R2017b (The MathWorks, Inc., Natick, MA). All EEGs were downsampled to 128 Hz, after which semi-automatic artifact detection was performed as described in(Lustenberger et al., 2015). Channels containing artifacts in more than 5% of all 20-sec epochs were excluded from further analysis. The recordings were then re-referenced to the average of O1 and O2, as these electrodes were available for all EEG recordings and slow wave activity (SWA) is typically most prominent in anterior regions in the age range of the investigated participants (Werth et al., 1997; Finelli et al., 2001; Kurth et al., 2010b, 2012).

A slow wave detection algorithm based on the method first described by (Riedner et al., 2007) was used for the detection of slow waves. EEG signals from all derivations were band-pass filtered (0.5 – 4.0 Hz, stopband 0.1 and 10 Hz) using a Chebyshev Type II filter (MATLAB, The MathWorks Inc, Natick, MA). Maximal attenuation in the passband was 3 dB and minimal attenuation in the stopband was 10 dB. Negative deflections of the EEG signal were depicted as ‘negative half-waves’. Negative half-waves with an interval of 0.25 to 1.0 seconds between the zero-crossings, i.e. wave frequency 0.5 – 2 Hz, were considered slow waves. These were detected in all artifact-free epochs of NREM sleep. Then, slow waves occurring in the first and last hour of NREM sleep were selected for slope analyzes, as has been performed in previous pathological and physiological pediatric data (Bölsterli et al., 2011; Fattinger et al., 2015; Kurth et al., 2016). Slopes of slow waves were calculated by dividing the amplitude of the most negative deflection by the interval until the next zero-crossing (Kurth et al., 2010a) (Fig. 1). Subsequently, they were corrected for amplitude as described in earlier work by calculating their value at the amplitude of 75 μV (further termed ‘slope’), because amplitude and slope are inherently correlated (Bölsterli et al., 2017, 2011; Bölsterli Heinzle et al., 2014). Furthermore, numbers of detected slow waves were determined for the FH of NREM sleep, LH of NREM sleep, and for the total time spent in NREM sleep, i.e. from the first to the last EEG epoch scored as NREM sleep. Mean slopes and numbers of detected slow waves were calculated for electrodes Fp1, Fp2, T3 or T5, and T4 or T6 (dependent on availability). In a second step, these values were averaged across these electrodes. The absolute and relative (i.e. with respect to the total sleep time) durations of sleep stages were calculated based on sleep stage-scored EEG epochs.

**Figure 1.**
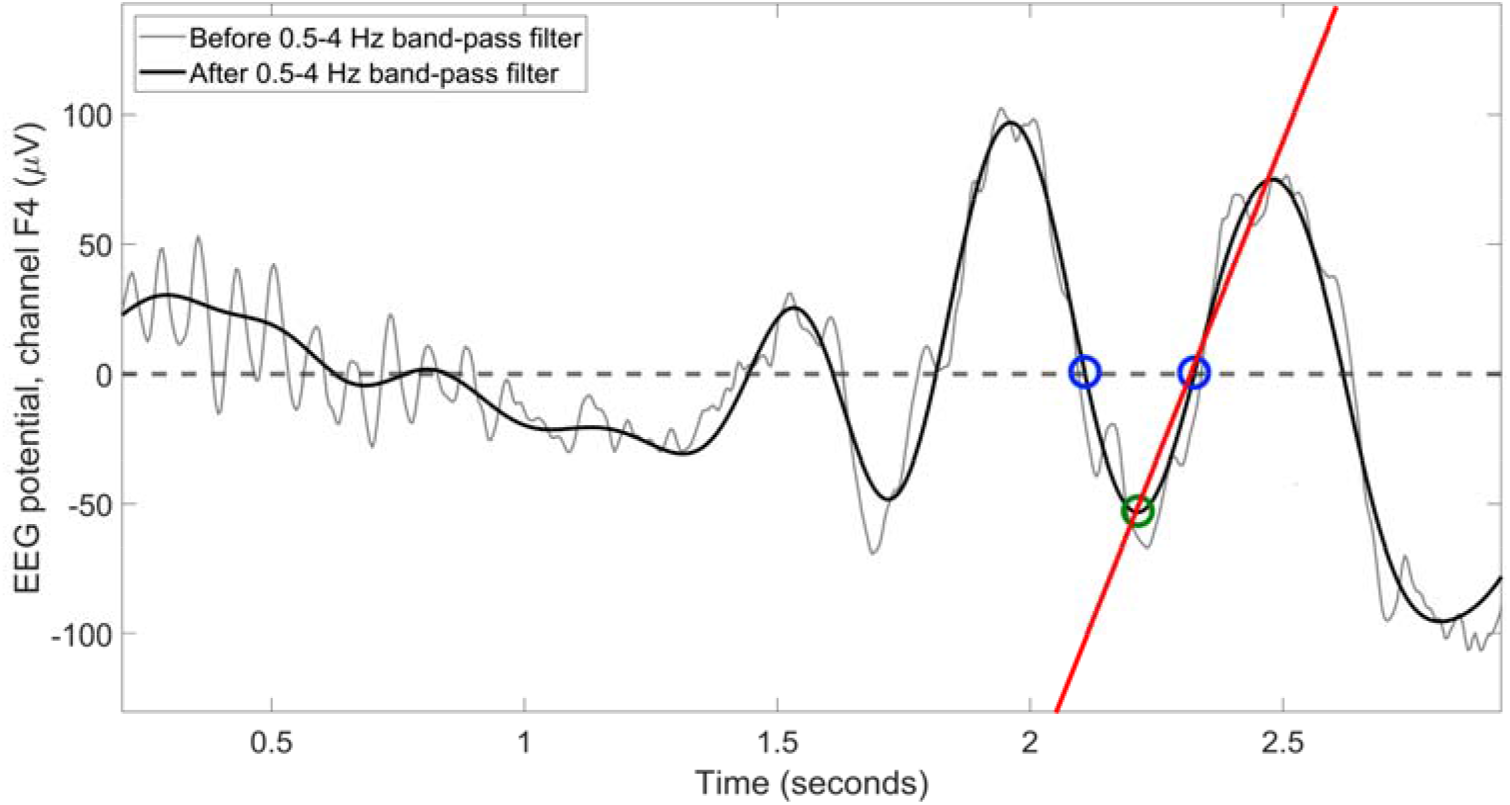
Example of a slow wave slope calculation demonstrated with an excerpt of the EEG of patient #5. The grey and black lines illustrate the EEG signal before and after application of the 0.5-4 Hz digital band-pass filter, respectively. Examples of detected zero-crossings of the band-pass filtered signal are depicted by blue circles, whereas the most negative value of the deflection in between is highlighted by a green circle. The slope of the corresponding slow wave was calculated based on the red straight line that passes through the minimum and the next zero-crossing.

### 2.4. Statistical analysis

Statistics were conducted with IBM SPSS Statistics (IBM Corp. Released 2017. IBM SPSS Statistics for Windows, Version 25.0. Armonk, NY: IBM Corp.). The one patient (#5) in whom the EEG was recorded clearly after clinical recovery, i.e. 4 years after the last manifestation of active disease, was excluded from quantitative statistical analysis. The corresponding slow wave analysis results were merely described qualitatively. In the statistical analysis, we included only those patient EEG recordings for which clinicians evaluated the hypnograms as normal, to eliminate influences of pathological sleep architecture on potential group differences in slow wave characteristics. This left 6 patients with 8 EEGs for statistical comparison to 30 controls. For each patient with two consecutive EEGs (designated EEG ‘A’ and ‘B’, respectively), ages at time of recording (2 to 10 weeks apart) and EEG analysis outcomes were averaged before any further analysis. Subsequently, statistical analysis was repeated, including also the patients and EEGs that showed pathologically fragmented sleep as described above. The results thereof are only reported in detail if they deviated from those previously reported.

For the analysis of slow wave slope variables between the two groups of participants, a 2×2 mixed ANOVA was performed with the within-subject factor ‘hour’ (FH vs. LH) and the between-subject factor ‘group’ (patients vs. controls). Standardized residuals were checked for the assumption of normal distribution (Shapiro-Wilk test). Following a significant interaction, we performed a priori defined post hoc tests, i.e. two-tailed independent samples *t*-tests, comparing slopes during the FH and the LH between patients and controls, with Bonferroni correction for multiple comparisons (realized by multiplying *p*-values by the number of post hoc comparisons). In case residuals of the mixed ANOVA were deviating from a normal distribution, pairwise non-parametric procedures (two-tailed Mann-Whitney *U*-tests) with Bonferroni correction for multiple comparisons were directly performed instead, comparing the slope changes between the FH and the LH, slopes during the FH, and slopes during the LH between patients and controls.

We compared further sleep EEG characteristics, i.e. numbers of detected slow waves and distribution of sleep stage distribution, directly between the groups. As some of the variables were not normally distributed, we applied non-parametric testing (two-tailed Mann-Whitney *U*-test) to all these variables.

Pearson correlation coefficients were calculated to assess the significance of associations between variables if residuals were normally distributed. Spearman correlation coefficients are reported otherwise. The threshold for significance for statistical analyzes was set at *p* < 0.05.

### 2.5. Data and code availability

MATLAB scripts used for the analyses in this study are provided by the corresponding author upon reasonable request.

## 3. Results

### 3.1. Changes of slow wave slopes across the night

In the analysis of slopes of slow waves during the FH and LH of NREM sleep in patients (#1 (B), #2, #4, #6, #7, and #9) and controls, we found a significant main effect for both hour (*F*(1, 34) = 33.31, *p* < 0.001, ANOVA) and group (*F*(1, 34) = 4.65, *p* = 0.038, ANOVA). This signifies that slopes were, in general, higher during the FH than during the LH (+54.02 ± 9.36 μV/s, difference of means ± standard error of difference, based on estimated marginal means) and overall higher in controls compared to patients (+43.02 ± 19.95 μV/s). Importantly, there was a significant interaction between hour and group (*F*(1, 34) = 6.91, *p* = 0.013, ANOVA). Namely, the decrease of the slope from the FH to the LH was significantly more pronounced in controls compared to patients (−49.21 ± 16.15 μV/s, Fig. 2). Post hoc tests revealed that controls showed significantly steeper slopes than patients during the FH (+67.62 ± 21.38 μV/s, *t*(34) = 2.598, *p* = 0.022, independent samples *t*-test), whereas there was no significant difference between the groups for slopes during the LH (18.41 ± 25.24 μV/s, *t*(34) = 1.074, *p* = 0.980, independent samples *t*-test, Fig. 2). Besides, while controls demonstrated a strong correlation between the slope during the FH of NREM sleep and the change of the slope across the night (*r*(28) = −0.845, *p* < 0.001, Pearson correlation coefficient, Fig. 3), there was no equivalent finding for patients with anti-NMDAR encephalitis (*r*(4) = 0.125, *p* = 0.814, Pearson correlation coefficient, Fig. 3). This means for control participants that, the steeper the slope was in the FH, the more it decreased towards the LH. In contrast, this relationship could not be confirmed in patients.

**Figure 2.** Slow wave slope for the first and last hour of non-rapid eye movement (NREM) sleep. Group mean values are provided for patients (black diamonds and dashed line) and control participants (grey circles and solid line). Error bars represent the standard error of the mean. The asterisk (*) indicates a significant interaction effect for ‘hour’ and ‘group’, i.e. the overnight (first hour to last hour of NREM sleep) change of the slow wave slope is different in patients and controls. Post hoc analyzes revealed a significant group difference for the first hour (marked by a black, solid triangle; *p* < 0.05; two-tailed independent samples *t*-test; Bonferroni corrected by multiplying the *p*-value by the number of post hoc tests), but not for the last hour of NREM sleep.

**Figure 3.** Slow wave slope during the first hour of non-rapid eye movement (NREM) sleep and the slow wave slope change across the night. Values are provided for patients (black diamonds) and control participants (grey circles).

Results were comparable when we included the three patient EEGs with abnormal sleep architecture (#1 (A), #3, and #8) in the analysis (data not shown).

### 3.2. Changes in individual participants

All controls consistently showed a decrease in the slope from the FH to the LH of NREM sleep (range −198.94 to −0.79 μV/s), whereas this was less consistent in patients (range −64.39 to 33.97 μV/s). Most of the latter showed a decrease (#1, #2, #3, #5, #6, and #9), but several also showed an increase (#4, #7, and #8), see Table 2 and Fig. 4. The overnight change of the slope in patient #5 (−31.57 μV/s), whose EEG was recorded 4 years after a relapse of anti-NMDAR encephalitis, was within the relatively small range of the values of controls in the corresponding age of a given patient (−25.43 to −55.96 μV/s) (Table 2, Fig. 4). Similarly, this patient’s slope values during the FH and LH of NREM sleep (363.63 μV/s and 332.05 μV/s, respectively) were either within or close to the values from the controls in the corresponding age (361.78 to 401.19 μV/s and 333.15 to 347.60 μV/s, respectively).

**Figure 4.**
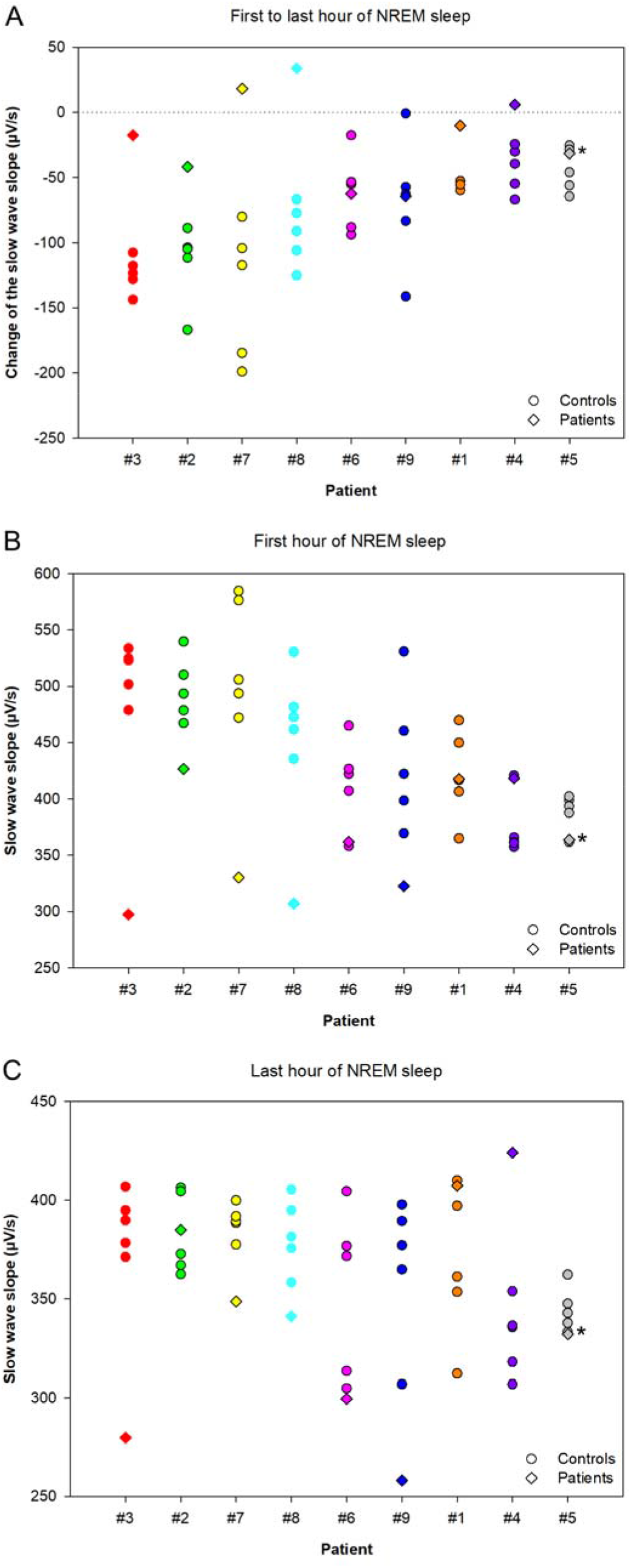
Slow wave slope variables for patients (colored diamonds) and for controls in the corresponding age of a particular patient (circles of the same color as the corresponding diamond). Participants are ordered according to their ages (increasing from left to right). Values are shown for the change of the slow wave slope across the night (panel A), the slow wave slope during the first hour of non-rapid eye movement (NREM) sleep (panel B), and the slow wave slope during the last hour of NREM sleep (panel C). Diamonds without black borders represent patients who had only EEGs with abnormal sleep architecture (see also 2. Materials and Methods) and accordingly, the healthy controls of corresponding age are indicated by circles without black lines. Patient #5, indicated with an asterisk (*), was measured in remission and was therefore not included in the analysis of patients during active anti-NMDAR encephalitis (see 2. Materials and Methods for details).

**Table 2.**
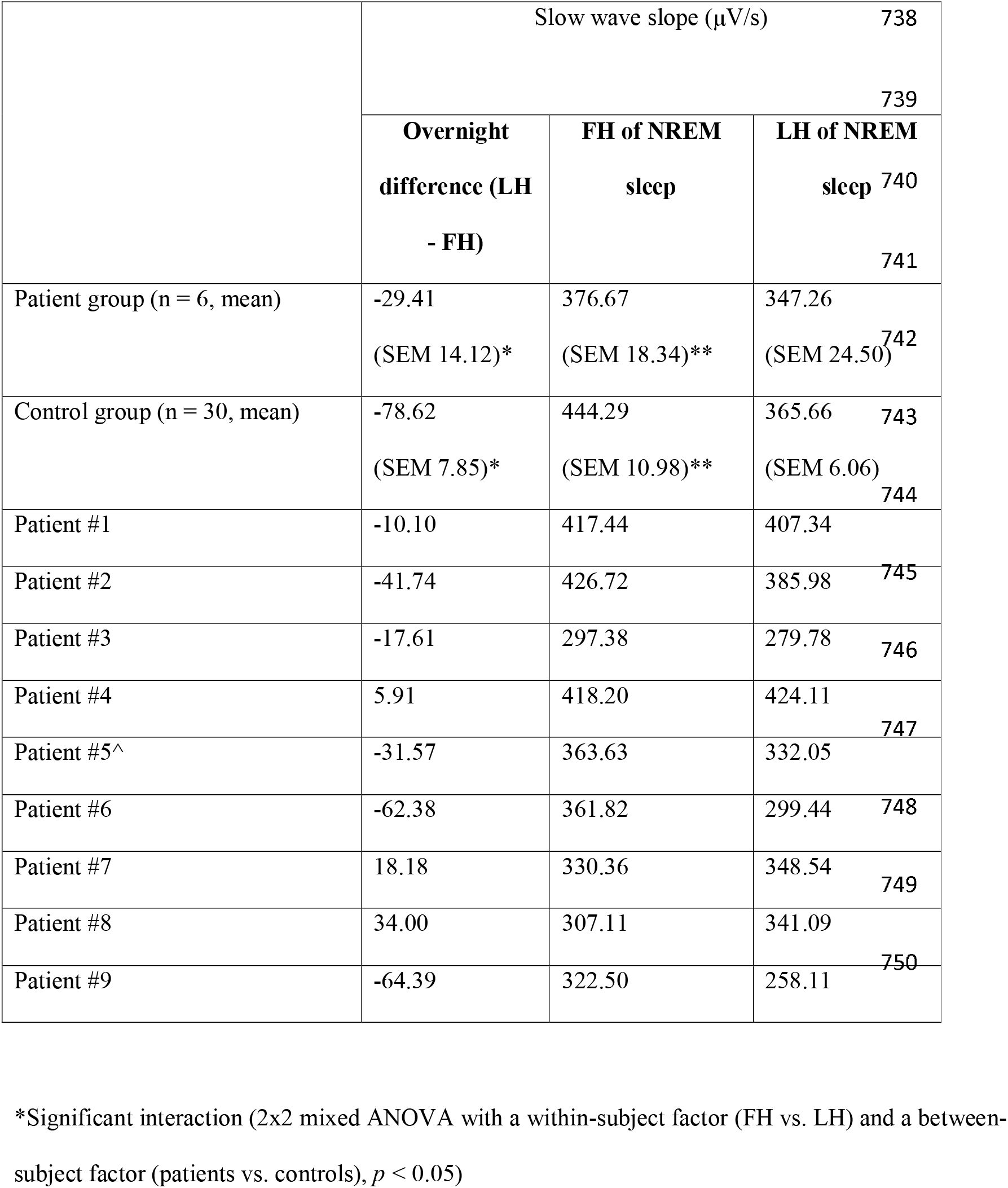
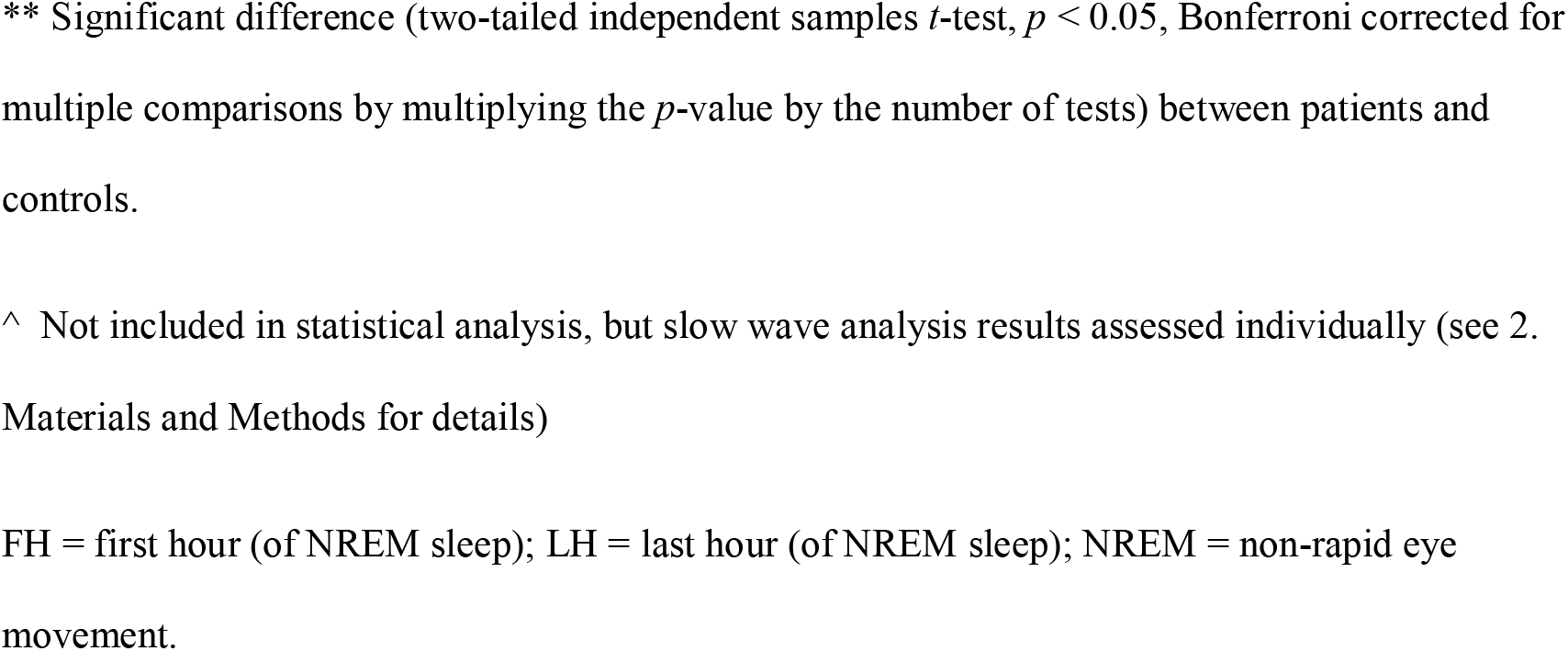
Slow wave slope values from (individual) patients with anti-NMDAR encephalitis and controls.

| | Slow wave slope (μV/s) | | |
| --- | --- | --- | --- |
|  | Overnight<br>difference (LH<br>- FH) | FH of NREM<br>sleep | LH of NREM<br>sleep |
| Patient group (n = 6, mean) | -29.41<br>(SEM 14.12)* | 376.67<br>(SEM 18.34)** | 347.26<br>(SEM 24.50)<sup>742</sup> |
| Control group (n = 30, mean) | -78.62<br>(SEM 7.85)* | 444.29<br>(SEM 10.98)** | 365.66<br>(SEM 6.06)<sup>743</sup> |
| Patient #1 | -10.10 | 417.44 | 407.34 |
| Patient #2 | -41.74 | 426.72 | 385.98<sup>745</sup> |
| Patient #3 | -17.61 | 297.38 | 279.78<sup>746</sup> |
| Patient #4 | 5.91 | 418.20 | 424.11<sup>747</sup> |
| Patient #5<sup>^</sup> | -31.57 | 363.63 | 332.05 |
| Patient #6 | -62.38 | 361.82 | 299.44<sup>748</sup> |
| Patient #7 | 18.18 | 330.36 | 348.54<sup>749</sup> |
| Patient #8 | 34.00 | 307.11 | 341.09<sup>750</sup> |
| Patient #9 | -64.39 | 322.50 | 258.11 |

\*Significant interaction (2x2 mixed ANOVA with a within-subject factor (FH vs. LH) and a between- subject factor (patients vs. controls), $p < 0.05$ )
\*\* Significant difference (two-tailed independent samples $t$ -test, $p < 0.05$ , Bonferroni corrected for multiple comparisons by multiplying the $p$ -value by the number of tests) between patients and controls.
^ Not included in statistical analysis, but slow wave analysis results assessed individually (see 2. Materials and Methods for details)
FH = first hour (of NREM sleep); LH = last hour (of NREM sleep); NREM = non-rapid eye movement.

### 3.3. Further sleep EEG characteristics

Numbers of detected slow waves during the FH of NREM, the LH of NREM, and all NREM of the sleep period were not different between the groups (Table 3). Repeating the analysis after including the three patient recordings with severe sleep disturbance yielded a significantly lower number of slow waves in patients (median 1584.56, IQR 1074.69, *U* = 78.00, *p* = 0.022, Mann-Whitney *U*-test) than in controls (median 2261.25, IQR 809.19) during the FH of NREM sleep. Yet, counts of slow waves were not different during the LH or all NREM sleep in the sleep period (data not shown).

**Table 3.** Further sleep EEG characteristics from patients with anti-NMDAR encephalitis and controls.

|  | Patients (n = 6) |  | Controls (n = 30) |  | *P*-value |  |
| --- | --- | --- | --- | --- | --- | --- |
|  | Slow wave count (#) |  | Slow wave count (#) |  |  |  |
| FH of NREM sleep | 1976 (1071) |  | 2425 (757) |  | 0.146 |  |
| LH of NREM sleep | 2099 (513) |  | 2189 (842) |  | 0.770 |  |
| Total NREM sleep | 10901 (4975) |  | 10153 (4894) |  | 0.544 |  |
|  | Absolute (hrs) | Relative (%) | Absolute (hrs) | Relative (%) | Absolute (hrs) | Relative (%) |
| Sleep period | 10.07 (1.54) | N.A. | 8.37 (1.48) | N.A. | <0.001* | N.A. |
| Wake (cont.) | 0.21 (0.58) | 2.26 (1.83) | 0.09 (0.31) | 1.11 (4.15) | 0.924 | 0.855 |
| Wake (cum.) | 0.64 (1.07) | 7.14 (8.81) | 0.43 (0.78) | 5.49 (7.66) | 0.733 | 0.823 |
| NREM 1 | 0.43 (1.62) | 4.46 (9.14) | 0.48 (0.38) | 5.57 (4.00) | 0.412 | 0.513 |
| NREM 2/3 | 5.74 (1.42) | 63.62 (27.26) | 5.49 (1.37) | 69.21 (13.31) | 0.667 | 0.564 |
| REM | 2.06 (1.32) | 21.39 (8.98) | 1.46 (0.84) | 18.27 (7.24) | 0.627 | 0.749 |

Variables are described by median and interquartile range (between parentheses). Two-tailed Mann-
Whitney *U*-tests were used in statistical comparisons between patients and controls.
\* Variables significantly different ( $p < 0.05$ ) between patients and controls.
cont. = continuous (longest period); cum. = cumulative; hrs = hours; N.A. = not applicable; NREM =
non-rapid eye movement (sleep); REM = rapid eye movement (sleep); Sleep period = time from the
beginning to the end of sleep (including periods of wakefulness).

In addition, patients did not significantly differ from controls in the length of any of the sleep-wake stages, apart from a longer sleep duration (time from falling asleep to waking up) during the EEG recording (difference of medians +1.73 hours, *U* = 15.00, *p* = 0.001, Mann-Whitney *U*-test) (Table 3). Comparable statistical outcomes were found when the three patient EEGs with disturbance of sleep architecture were included for analysis (data not shown).

### 3.4. Participants’ ages and changes of slopes across the night

Healthy individuals showed smaller overnight decreases of slow wave slopes with increasing age (Spearman’s rho(28) = 0.724, *p* < 0.001, Fig. 5). Contrastingly, such an age-related effect was not evident in patients (Spearman’s rho(4) = 0.086, *p* = 0.872). Results were comparable when the three initially excluded patient EEGs were added to the analysis (Fig. 5).

**Figure 5.** Participants’ age at the time of the EEG recording and change of the slope across the night. Patient values are shown by uniquely colored diamonds, whereas values from control participants in the corresponding age of a particular patient are represented by circles of the same color as the corresponding diamond. Diamonds without a black line denote EEGs of which hypnograms were determined by a clinician to demonstrate abnormal sleep architecture (see 2. Materials and Methods for a specification), with corresponding circles lacking a black border. Note that patient #1 had one EEG with abnormal sleep architecture (indicated by a diamond without black border) and one EEG with normal sleep architecture (indicated by a diamond with black border). Accordingly, the circles representing control EEGs have black borders in this case. As the residuals of the variables plotted were not normally distributed, Spearman correlation coefficients were calculated. In the figure, the linear regression line for patients (black, dashed) is for illustrative purposes only.

## 4. Discussion

In this study, we hypothesized that changes of slow wave slopes from the FH to the LH of NREM sleep, an indirect measure of synaptic plasticity (Riedner et al., 2007; Vyazovskiy et al., 2009), are altered in a human model of NMDAR deficiency caused by anti-NMDAR antibodies, i.e. patients with anti-NMDAR encephalitis. Results confirmed that the overnight decrease of the slope is significantly smaller for patients than for healthy controls, accompanied by significantly smaller slopes during the FH of NREM sleep. Contrastingly, the slope during the LH of NREM sleep does not significantly differ between patients and controls. These results are in support of our hypothesis and, therefore, suggest that the analyzed sleep EEG markers for synaptic plasticity are sensitive to an NMDAR deficiency as evident in anti-NMDAR encephalitis. We conclude that synaptic plasticity seems altered in this human model of NMDAR deficiency.

Various studies have proposed the functional involvement of cortical NMDAR in slow wave sleep (NREM stage 3) (Armstrong-James and Fox, 1988; Bazhenov et al., 2002; Duncan et al., 2013). Yet, disruption of NMDAR function in animals revealed that these receptors are additionally engaged in changes of EEG slow waves following synaptic potentiation during wakefulness (Campbell and Feinberg, 1996; Miyamoto et al., 2003). Electro-encephalographic abnormalities have also been investigated in anti-NMDAR encephalitis patients, i.e. human models of NMDAR deficiency (Benjumea-Cuartas et al., 2017; Gitiaux et al., 2013; Nosadini et al., 2015). However, to the best of our knowledge, this is the first study comparing slow wave characteristics during NREM sleep between patients with anti-NMDAR encephalitis and healthy individuals.

The synaptic homeostasis hypothesis may contribute to the understanding of the observations in patients with functional NMDAR deficiency. It states that the degree of net synaptic downscaling during sleep is closely tied to the net increase in synaptic strength during prior wakefulness (Tononi and Cirelli, 2014). Synaptic strengthening during wakefulness is the result of long-term potentiation (LTP) of cortical circuits, which is assumed to be the primary mechanism of learning (Dupuis et al., 2014). Numerous studies suggested that the synaptic strength of cortical connections is reflected by the slope of slow waves (Esser et al., 2007; Riedner et al., 2007; Vyazovskiy et al., 2009). As NMDAR are crucial for the induction of LTP (Collingridge and Bliss, 1987), the smaller slopes during the FH of NREM sleep in patients could indicate less LTP during wakefulness because of NMDAR deficiency.

However, NMDAR also play a role in promoting long-term depression (LTD) of synaptic networks (González-Rueda et al., 2018; Nabavi et al., 2013). LTD is predominant during sleep, causing a net decrease of synaptic strength (Tononi and Cirelli, 2014). The smaller slope decrease in patients with NMDAR deficiency may, therefore, indicate that also LTD is impaired. As patients were at different stages of their disease, different degrees of NMDAR deficiency and thus alterations in synaptic plasticity were expected, which is supported by the high variability of overnight changes of slopes in patients (Table 2, Fig. 4). Moreover, to achieve homeostasis of synaptic strength, we would expect these overnight changes of slopes to relate to the level at the beginning of sleep resulting from previous wakefulness. The lack of a correlation between slopes during the FH of NREM sleep and their change during sleep in patients (Fig. 3) implies that the homeostatic regulation of slow waves is disrupted. This indicates a dysfunction of synaptic homeostasis independent of smaller slopes at the beginning of sleep.

Functional NMDAR are considered crucial in the maintenance of normal sleep patterns (Tomita et al., 2015; Vataev et al., 2014). Aberrant sleep patterns, observed in patients with anti-NMDAR encephalitis (Florance et al., 2009; Gitiaux et al., 2013; Irani et al., 2010), may theoretically influence the overnight changes of slow waves. To minimize such a confounding effect, only patient EEGs with visually normal appearing hypnograms were included in the main statistical analysis. Besides longer total sleep periods in patients, neither durations and proportions of sleep stages, nor the numbers of detected slow waves differed between patients and controls (Table 3). Additionally, by including controls with similar age as patients, we prevented that age-related differences in overnight slope decrease – as confirmed to exist in healthy subjects (Fig. 5) – would have driven the outcomes. Therefore, the altered slow wave slope values in our patients are likely to be the result of factors beyond altered sleep architecture and age.

The majority of patients in this study were female (7/9), corresponding with the fact that females are predisposed for the development of anti-NMDAR encephalitis (Suh-Lailam et al., 2013). One of them, patient #5, was measured more than 5 years after diagnosis and about 4 years after a relapse of the disease. She had no clinical sequelae. Clinical recovery from anti-NMDAR encephalitis occurs due to a decrease of anti-NMDAR antibodies, leading to the recuperation of NMDAR function (Hughes et al., 2010). Recuperated NMDAR function could be explanatory for results from patient #5, being within or close to the narrow range of values of controls in the corresponding age of a given patient (Fig. 4). However, since we do not have an EEG during active disease, it remains unknown whether slow wave slope variables were altered at that time. Reversibility of pathologically altered overnight changes of slow wave slopes has been shown in ESES (Bölsterli et al., 2017), a disease that has been linked to a genetically determined gain of NMDAR function (Carvill et al., 2013; Lemke et al., 2013; Lesca et al., 2013).

This study also has limitations. One of them is certainly the small number of patients included (n = 9). However, given the rarity of pediatric anti-NMDAR encephalitis (Leypoldt et al., 2013) and the scarcity of whole-night EEGs in its patients, we believe the size of our dataset is reasonably large. In particular, the large control group with the same age distribution rendered it unlikely that differences arose from the selected controls since it accounted for the variance that exists between individuals in a specific age range. Additionally, 9/21 EEGs were not analyzable due to artifacts and severely altered background activity preventing sleep stage scoring. Patients in the acute phase of the disease are frequently agitated and suffer hyper-motor movement disorders; both reasons for EEG artifacts. These issues may have introduced a selection bias towards patients less affected at the time of the EEG. This possible bias would reduce differences compared to healthy controls. Importantly, despite the small sample size and EEGs of rather less affected individuals, we nonetheless demonstrate significant group effects.

Another limitation of this retrospective study is that it is unknown whether participants slept during the day before the night of the EEG recording. For those who did, slopes in the FH of NREM sleep during the EEG recording may have been lowered by both a reduced time of synaptic potentiation during wakefulness and a decrease of synaptic strength during this daytime sleep. Moreover, lacking information on the occurrence of seizures in patients the day preceding their whole-night EEGs could have additionally systematically impacted the results. However, since seizures have been shown to be related to increased SWA and slope of slow waves in the sleep EEG (Boly et al., 2017), one would expect an increase of slope of slow waves in the sleep period after wake-time seizures. As this would have weakened our results, the differences between patients and controls would be even stronger when accounting for this.

A possible confounder could have been that most of the patients with active disease underwent drug treatment at the time of the EEG registration. This included corticosteroids and a variety of anti-seizure agents (Supplementary Table 1). Various substances have been suggested to alter slow wave sleep in different directions (Holsboer et al., 1988; Legros and Bazil, 2003). In earlier studies of our group, we showed that corticosteroids reduce the slope of slow waves in infants with infantile spasms. However, the overnight change of slope was not investigated (Fattinger et al., 2015). Due to the small sample size in this study, the effect of corticosteroids could only be analyzed qualitatively. Slope values during the FH and LH, as well as overnight changes of the slope in patients on corticosteroids, were below, within, and above the levels of controls (Fig. 4). Hence, a systematic influence of corticosteroid treatment on how slope values differed from the controls in the corresponding age of a given patient is highly unlikely. Additionally, the wide variety of anti-seizure drugs hindered grouping of patients to accurately estimate their effects. Consequently, we cannot exclude an influence of anti-seizure drug treatments on our results, even though a systematic effect is improbable because of the large variability in treatments. Despite these limitations, we believe that our study is unique as it contains a group of young patients with a very rare, yet well-defined disease (by the occurrence of clinical symptoms and anti-NMDAR antibodies), which we compared to a large healthy control group of the same age and (nearly) same sex distribution.

Future perspectives related to this first study on overnight changes of slow wave slopes in a human model of NMDAR deficiency would be to conduct prospective and ideally longitudinal multicenter studies with standardized data acquisition (clinical, laboratory, imaging and EEG data) in larger samples of patients with anti-NMDAR encephalitis. Such studies could clarify whether dysregulated changes of slow wave slopes across the night can account for clinical symptoms and if recovery of overnight changes of the slopes is accompanied by a clinical recovery in patients. Alternatively, NMDAR-related parameters could be estimated by the use of generative models such as dynamic causal models (DCM), an approach that has been applied in previous studies in anti-NMDAR encephalitis (Cooray et al., 2015; Rosch et al., 2018; Symmonds et al., 2018), but not with regard to sleep EEG characteristics. Besides, more direct markers of functional NMDAR, for example levels of autoantibodies against NMDAR, could be related to the EEG markers of synaptic plasticity. This would be a next step towards a novel, clinically applicable, electrophysiological biomarker for pathologies of synaptic plasticity.

In conclusion, patients with anti-NMDAR encephalitis compared to controls show a smaller decrease of slow wave slopes over the course of the night and smaller slopes at the beginning of the night. As these patients are known to have reduced functional NMDAR and as changes in slow wave slopes have been suggested to reflect synaptic plasticity, our results support the presumption that synaptic plasticity during sleep and wakefulness, including synaptic homeostasis, is hampered due to functional NMDAR deficiency. In future, the slope of slow waves might be used as an instrument for recognition of altered neuronal plasticity, for example due to altered NMDAR function in diseases and physiological situations such as learning or normal development.

## Acknowledgements

This work was supported by radiz - Rare Disease Initiative Zurich (to S.R.G.), the Swiss National Science Foundation (PCEFP1-181279) (to S.K.), the Swiss National Science Foundation (320030-179443 and 320030-153387) (to R.H.), the Clinical Research Priority Program “Sleep and Health” of the University of Zurich (to R.H.), the Anna Müller Grocholski foundation and the EMDO foundation (to B.K.B.).

We thank Jakob Heinzle for his suggestions and inputs regarding the statistical approach.

## Supplementary material

**Supplementary Table 1.**
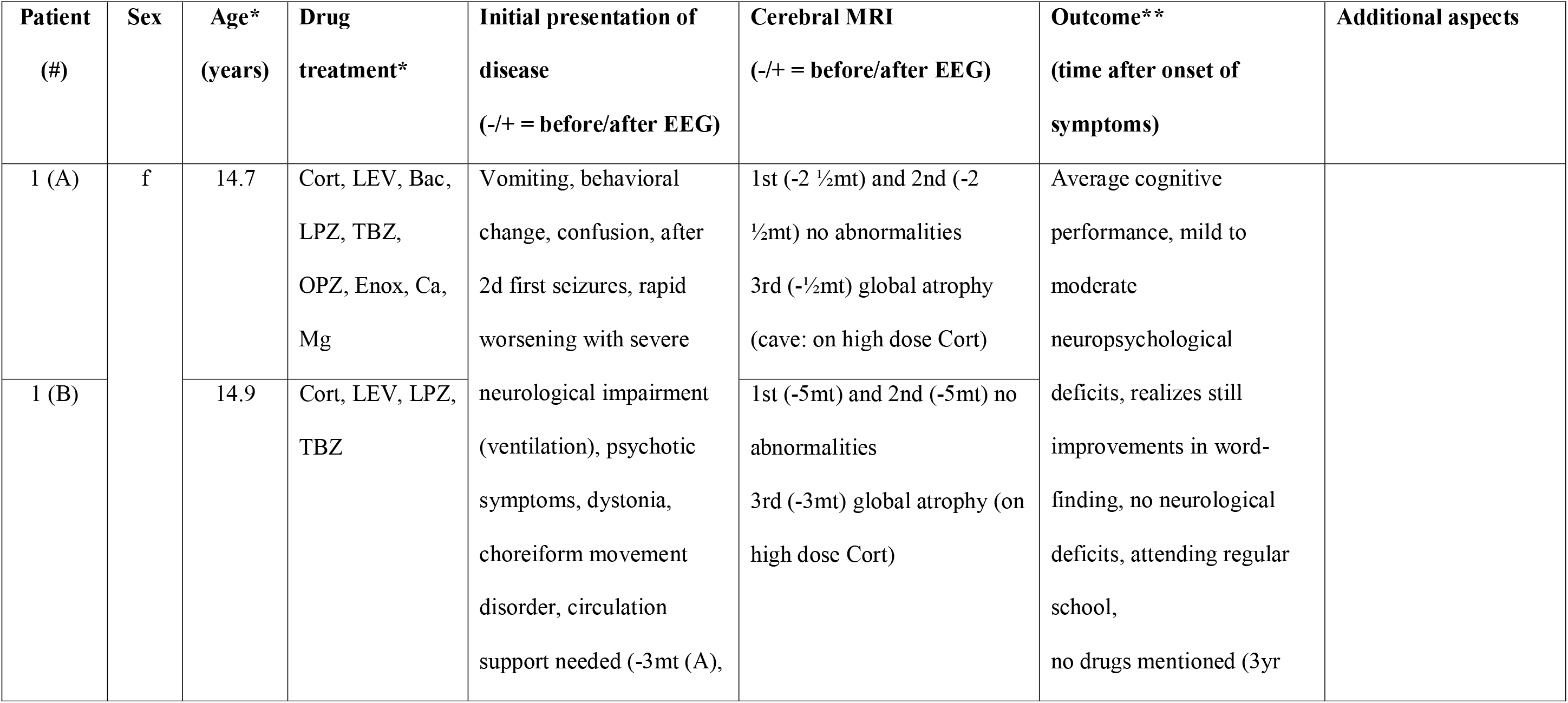
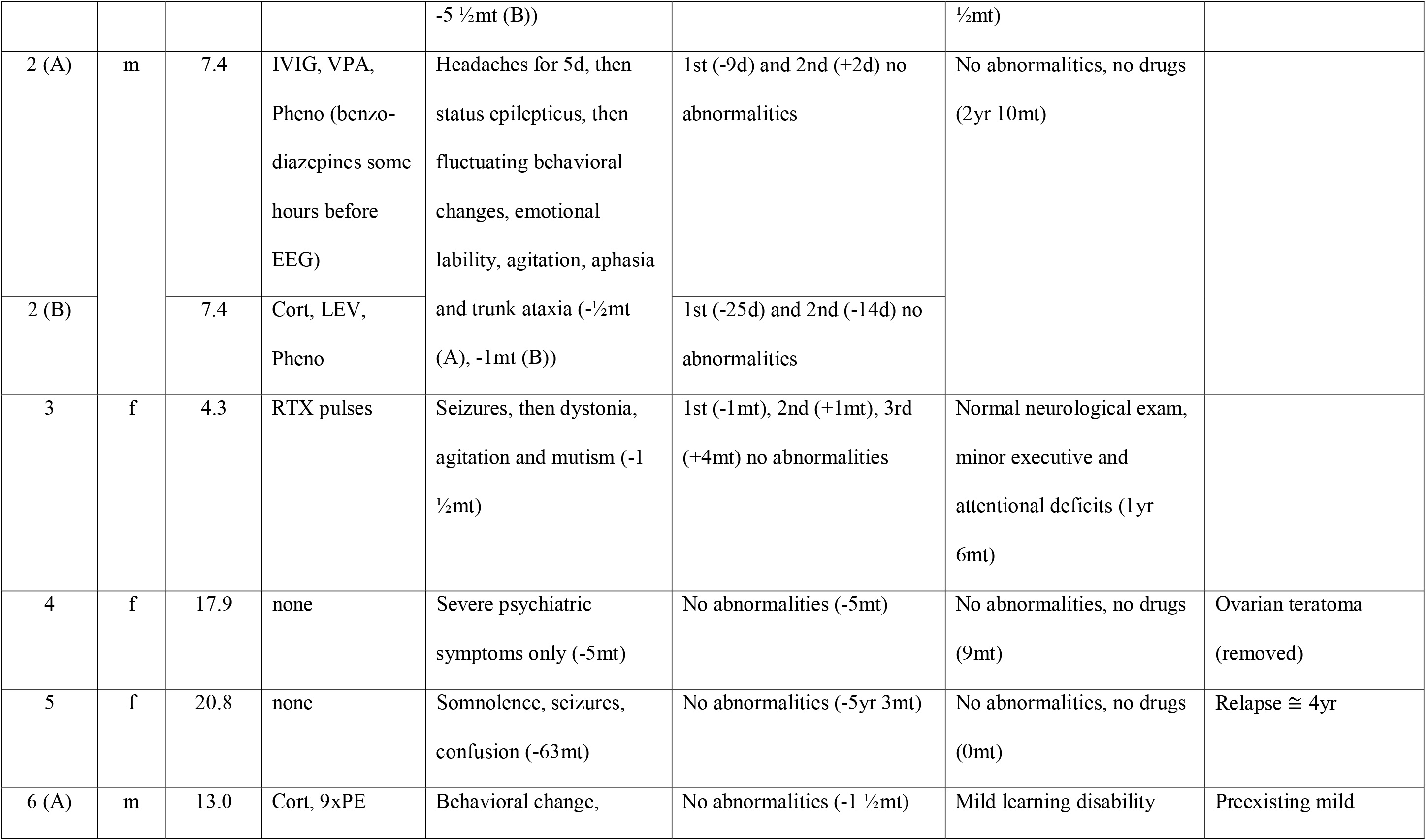
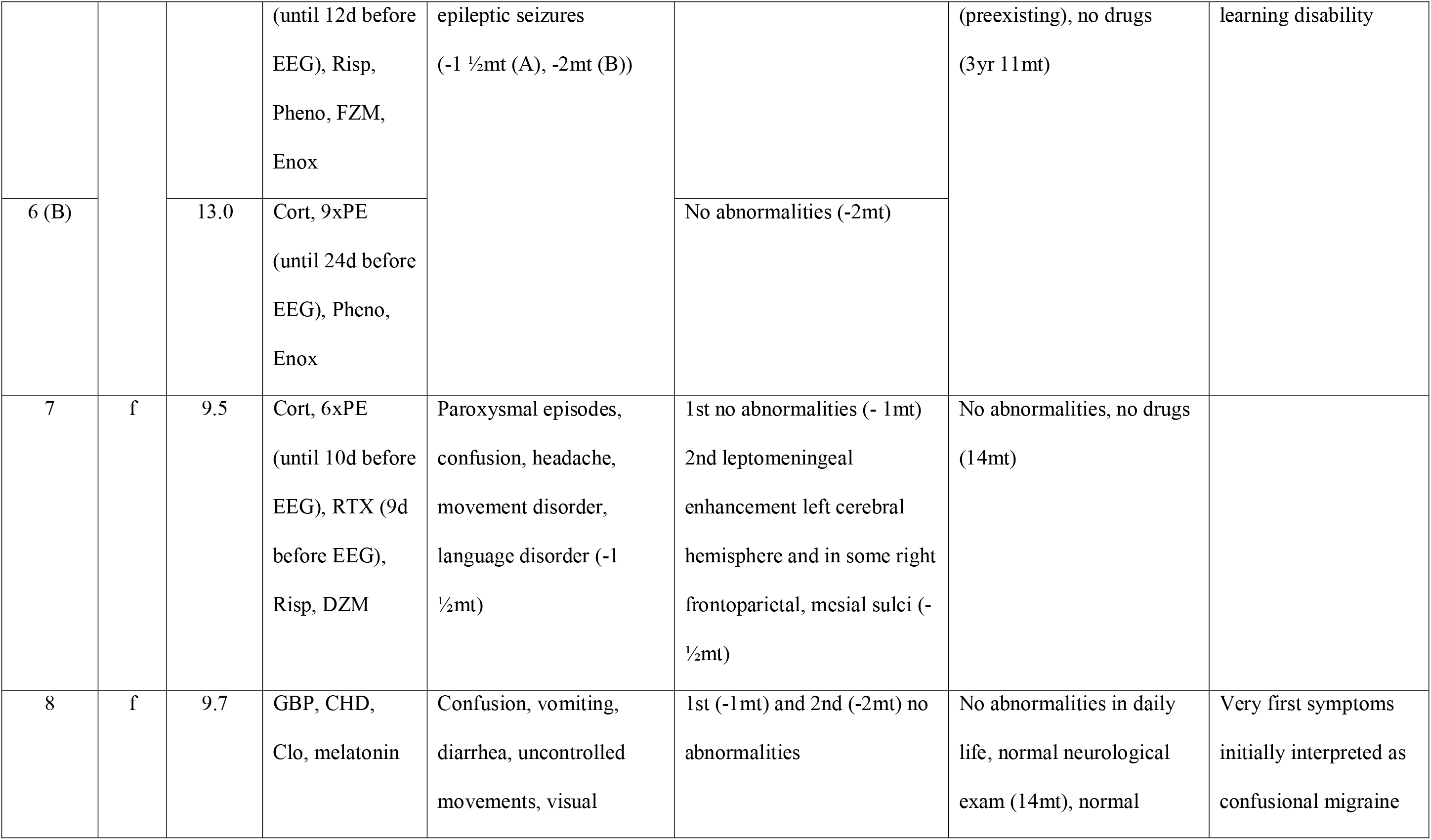
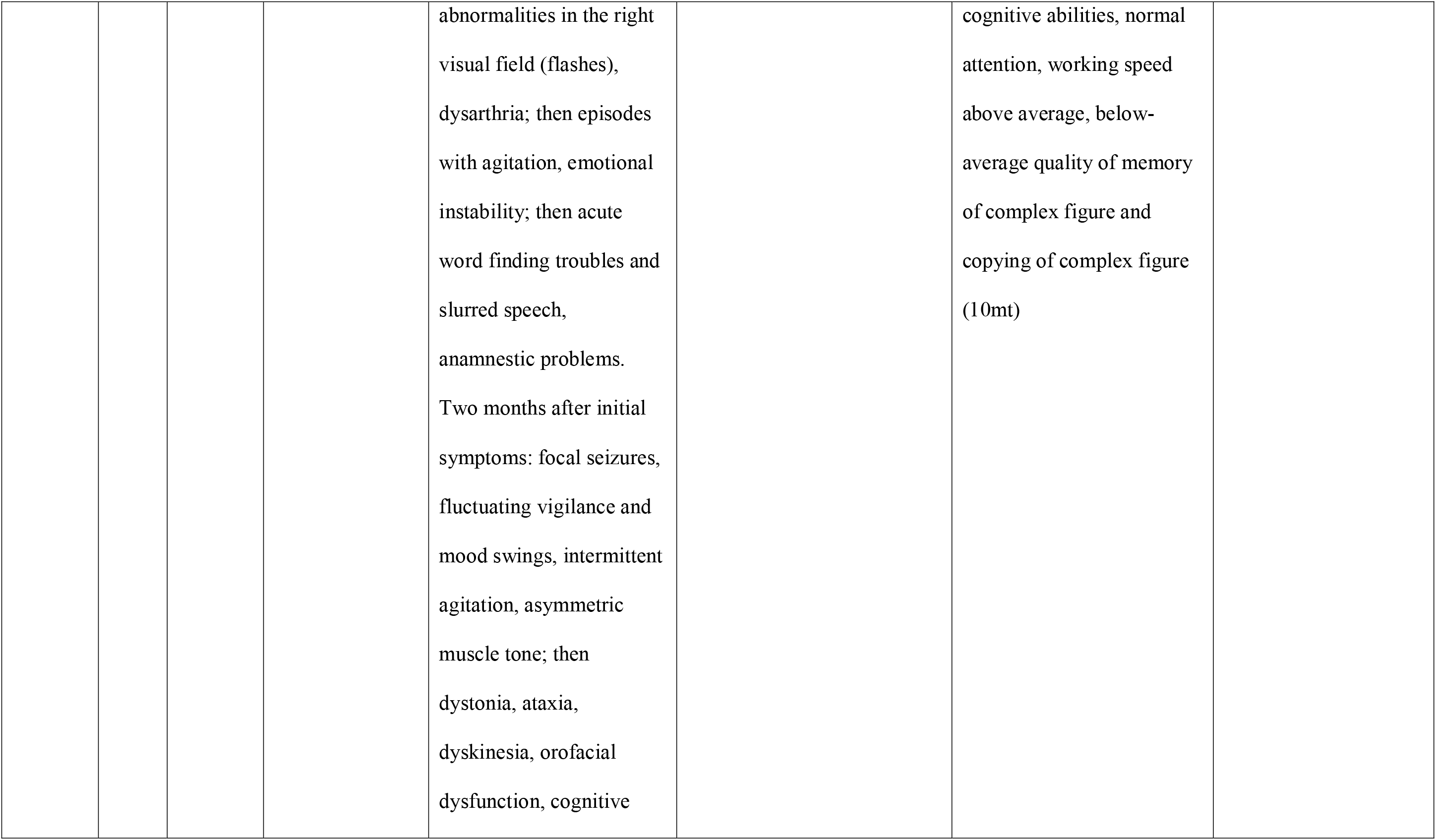
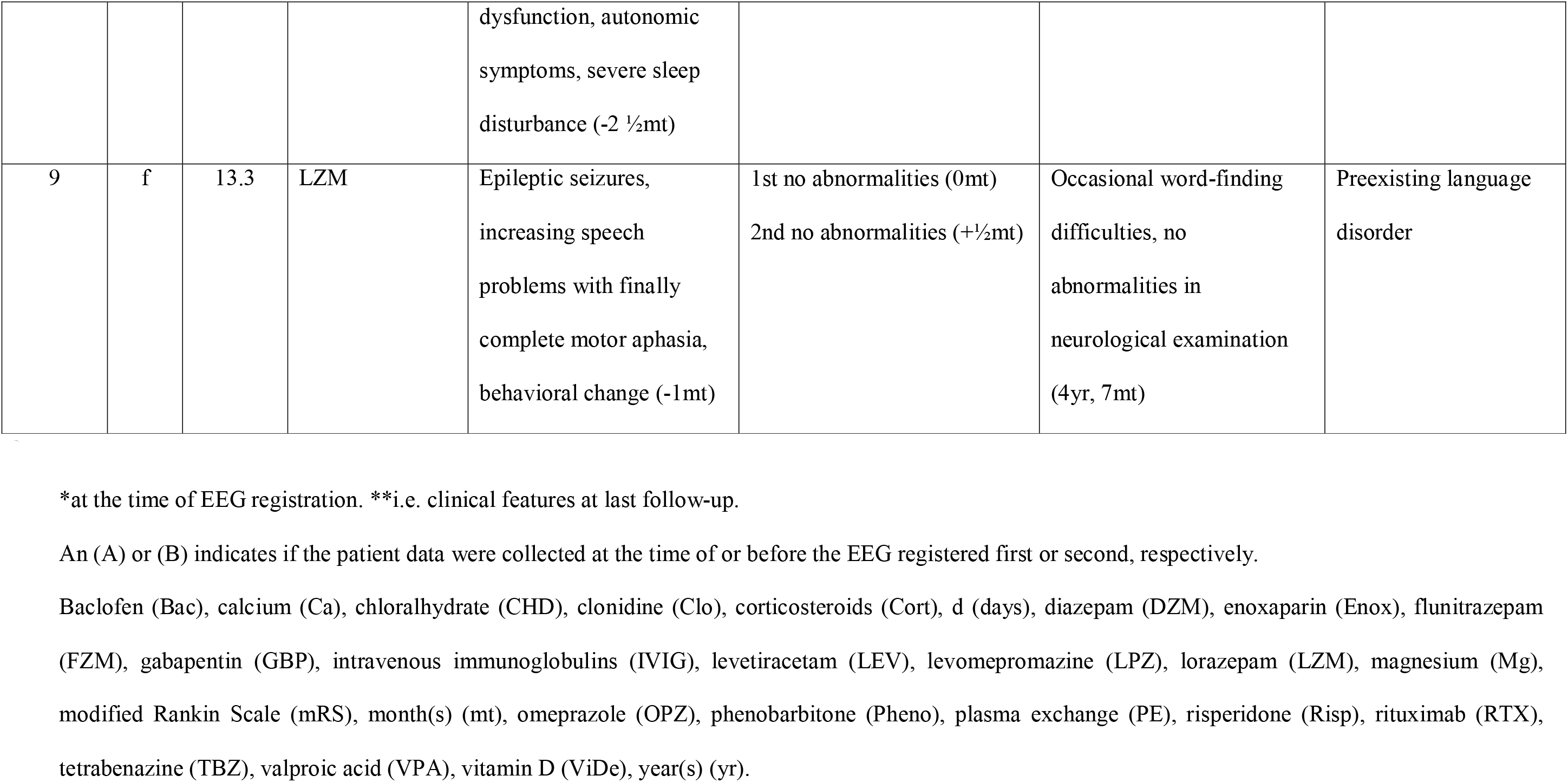
Additional clinical characteristics of patients with anti-NMDAR encephalitis.

### Pediatric mRS

mRS 0 No symptoms at all

mRS 1 No significant disabilities despite symptoms in clinical examination; age-appropriate behaviour and further development

mRS 2 Slight disability; unable to carry out all previous activities, but same independence as other age- and sex-matched children (no reduction of levels on the gross motor function scale)

mRS 3 Moderate disability; requiring some help, but able to walk without assistance; in younger patients, adequate motor development despite mild functional impairment (reduction of one level on gross motor function scale)

mRS 4 Moderately severe disability; unable to walk without assistance; in younger patients reduction of at least 2 levels on the gross motor function scale

mRS 5 Severe disability; bedridden, requiring constant nursing care and attention

mRS 6 Dead

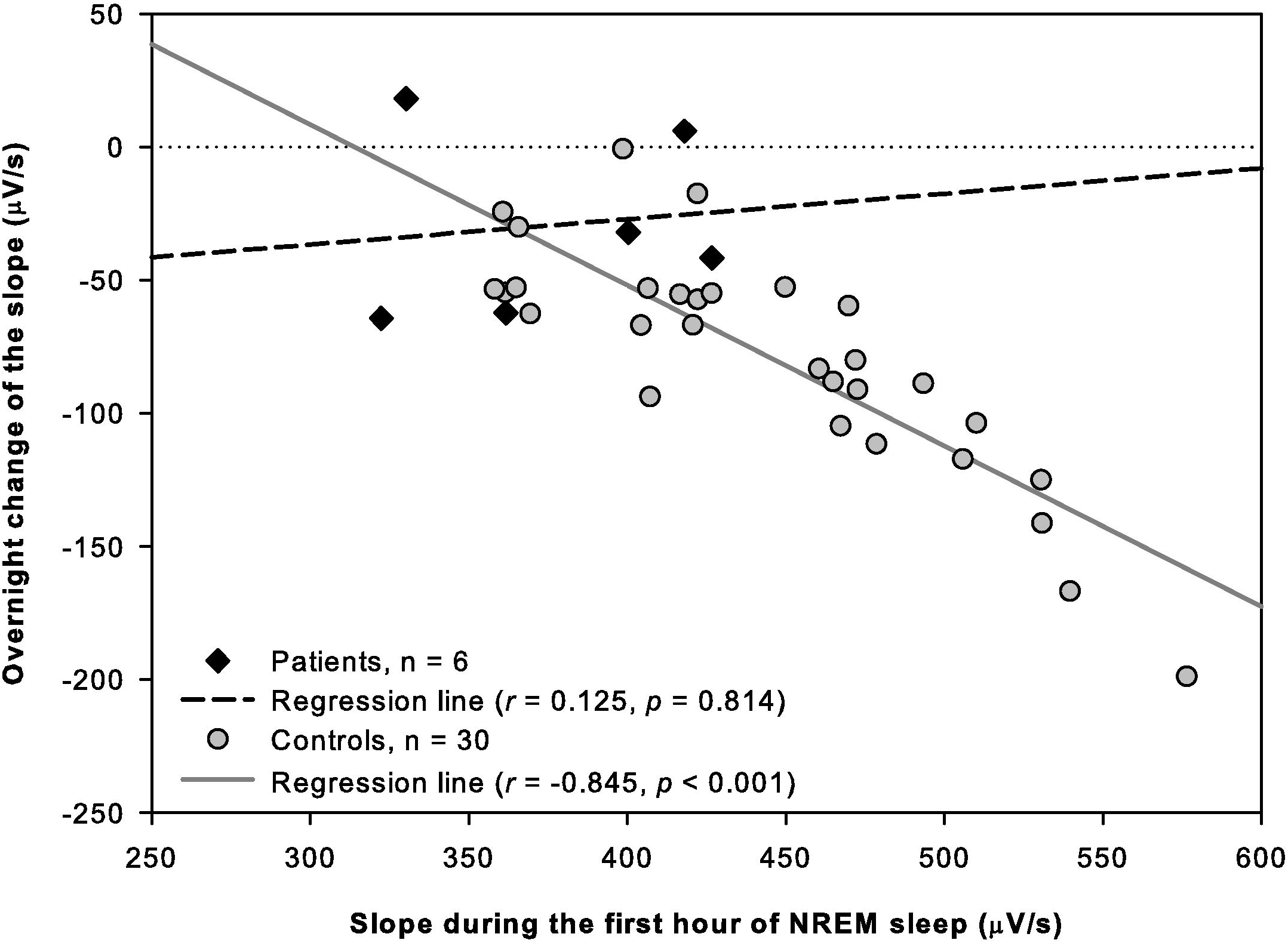

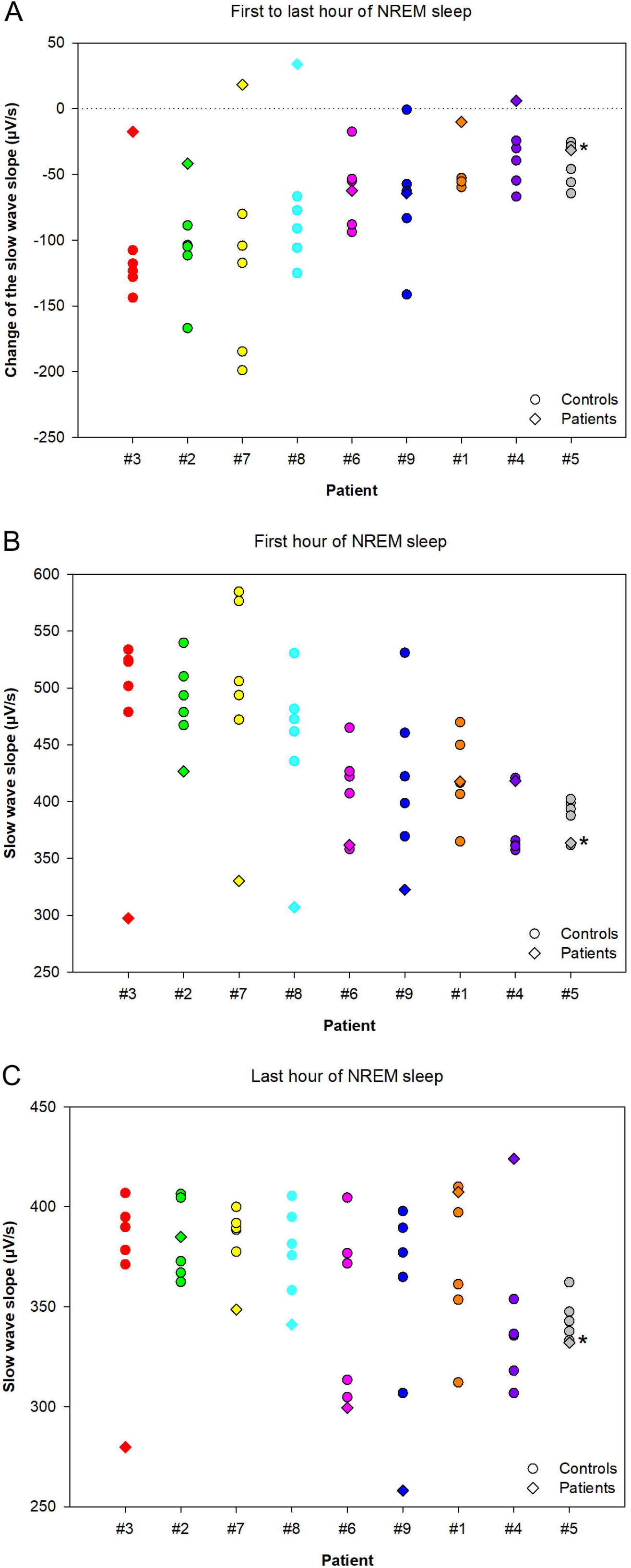

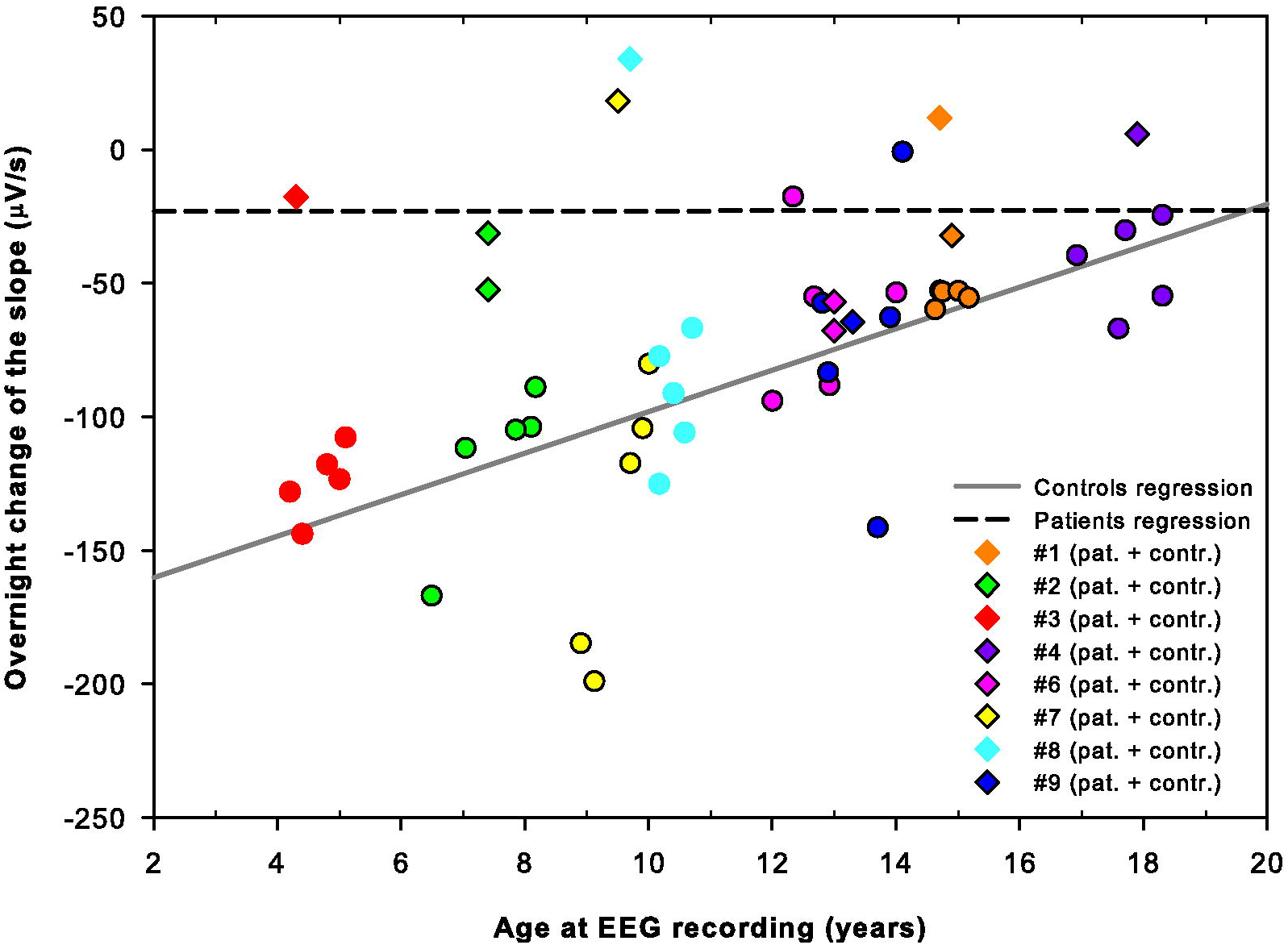

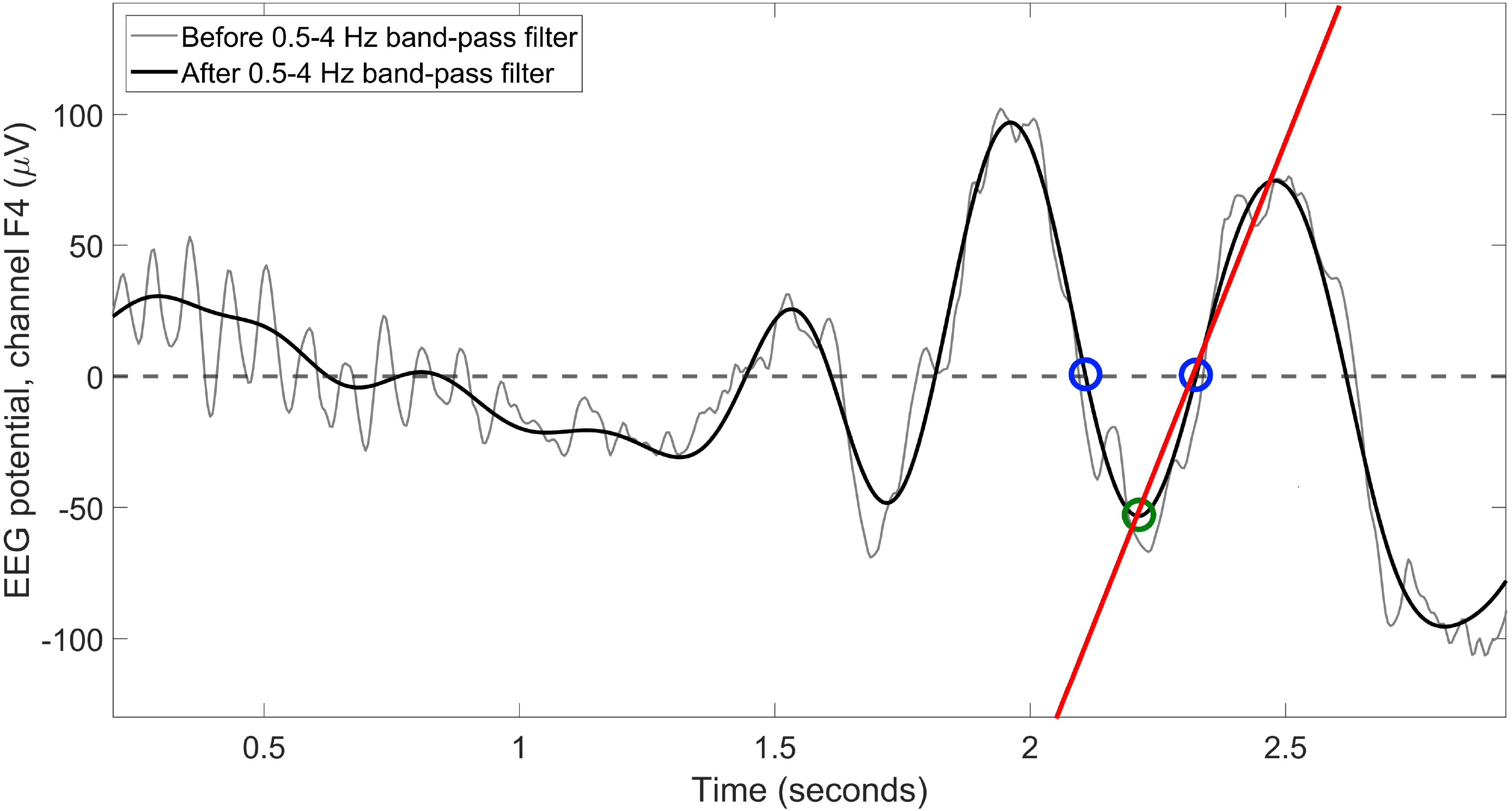

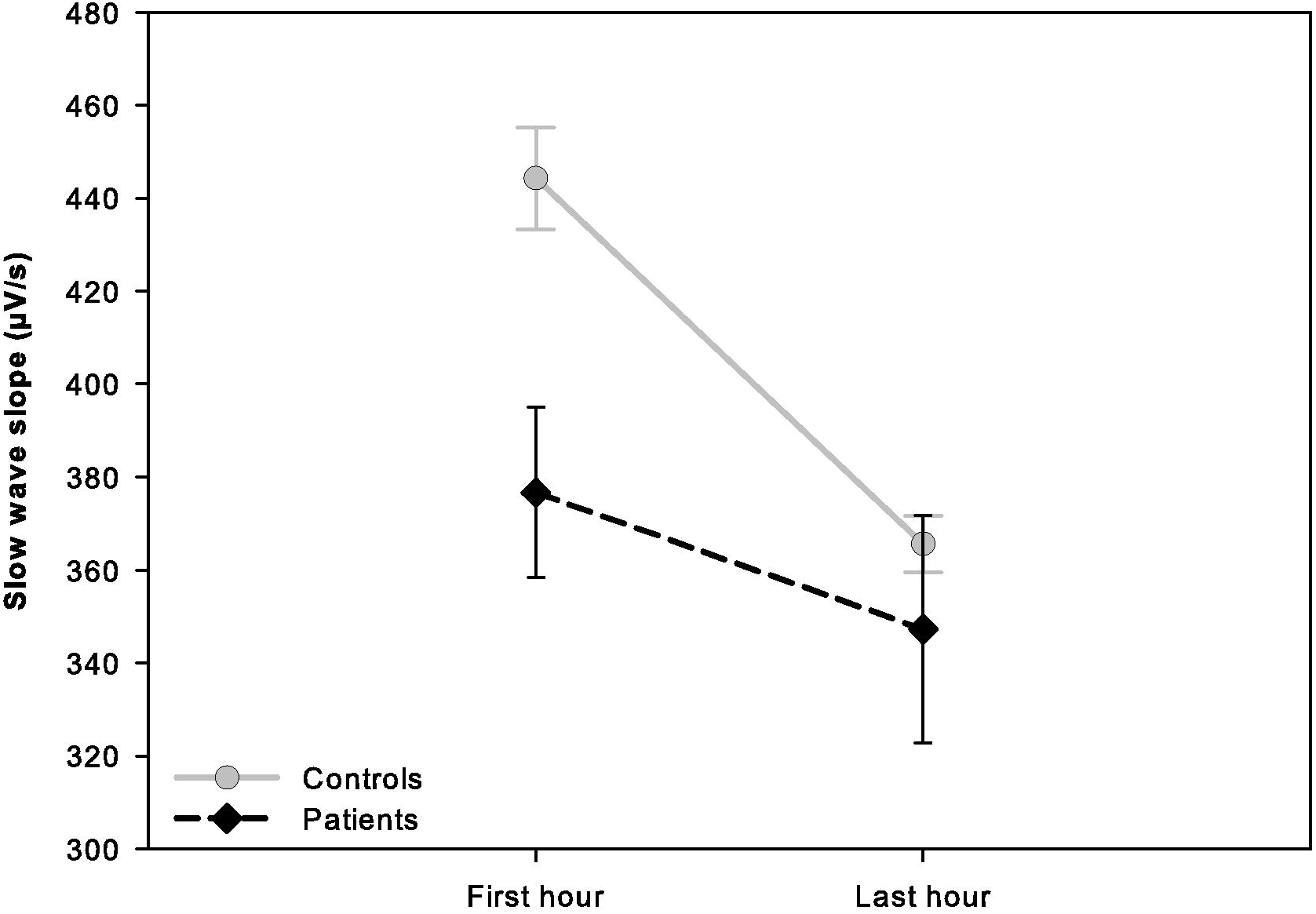

